# Registration and Reporting of Clinical Trials Affiliated with California Universities and with Primary Completion Date from 2014 to 2017

**DOI:** 10.1101/2025.04.07.25325421

**Authors:** Mario Malički, Vladislav Nachev, Susanne Wieschowski, Nicole Hildebrand, Stefanie Gestrich, Samruddhi Yerunkar, Emmanuel Zavalis, Benjamin Gregory Carlisle, Delwen L. Franzen, Maia Salholz-Hillel, Steven N. Goodman, Daniel Strech

## Abstract

**Background:** Public information on US clinical trials is shared through the ClinicalTrials.gov registry. This study’s goal was to determine prospective registration, results reporting, trial registration number reporting, and publication accessibility status for trials with primary completion dates from 2014 to 2017 affiliated with seven California universities.

**Methods:** We identified trials with investigators, sponsors, or responsible parties affiliated with seven California universities and searched for their results publications manually. We then used semi-automatic methods to determine prospective registration, summary results reporting in a registry, publication status, and reporting of registration numbers in the abstract and the full text of manuscripts.

**Results:** We identified 1,091 unique trials. Most trials were single-center (n=752, 69%) and had a median of 50 participants (IQR 21 to 150). Overall, 64% of trials (n=698) were prospectively registered, 46% (n=500) had summary results reported in the registry, 69% (n=750) had results published as articles, and an additional 3% (n=36) as abstracts or posters. Results reporting (summary, articles, abstracts or posters) occurred for 58% of trials (n=637) within 2 years, and for 81% (n=888) within 5 years of study primary completion date. Of journal publications, 77% (n=579) were open access publications, 37% (n=276) had trial registration numbers listed in their abstract, and 45% (n=336) in the manuscript. Only 92 (8%) of these trials were legally required to report results, and only 2 (2%) of those were overdue and under primary responsibility of a California university to report.

**Conclusions:** Almost one fifth of clinical trials with primary completion dates from 2014 to 2017 with investigators, sponsors or responsible parties affiliated with seven California biomedical research universities lacked any results reporting 5 years after their primary completion, and only 58% reported results within 2 years. Even though a large majority of these trials were completed before US legal mandates for reporting, there was an ethical requirement that the burden to research participants should be commensurate with the scientific value of the research. Research has no public scientific value if its results are not reported.

## Introduction

For clinical trials to generate useful and unbiased medical knowledge, trials should be registered before they start, and results should be reported after they are completed. These principles have been part of the Declaration of Helsinki since 2008 regarding registration and since 2013 regarding reporting.(1) More recently, this general guidance for publication of results has been specified to mean that results reporting should be timely, defined as summary results reporting in a registry within 1 year of primary completion date, and results publication in peer-reviewed journals within 2 years.(2) These ethical recommendations are relevant regardless of legal requirements.

The ClinicalTrials.gov trial registry was launched in 2000, and in 2008 it expanded to include summary results reporting.(3) The Food and Drug Administration Amendments Act of 2007 (FDAAA), and the later 2016 Final Rule for Clinical Trials Registration and Results Information Submission, applicable to specific US trials, makes US trial registration mandatory within 21 days of first patient enrollment and summary results reporting within 12 months of trial primary completion date. Similarly, the National Institutes of Health (NIH) 2016 Policy on the Dissemination of NIH-Funded Clinical Trial Information (effective from January 18, 2017) requires the same for clinical trials funded in whole or in part by NIH. Information in the ClinicalTrials.gov registry is self-reported by trial sponsors or investigators through a web-based Protocol Registration and Results System (PRS), with each entry having a set of mandatory data elements that are reviewed by the ClinicalTrials.gov staff. Each entry is also expected to be updated throughout the trial’s life cycle, with all changes made visible through the “History of Changes” links on the registry website.(3)

Studies have shown that while trial registries are the only publicly accessible source of results information for thousands of trials,(4) many trials are not prospectively registered,(5) trials can have discrepancies between outcomes listed in their registration and their peer-reviewed publications, and not all trials have summary results reported.(6–8) Because of the latter, initiatives like the FDAAA TrialsTracker publicly track compliance.(9) In April 2025, according to TrialsTracker, 78% (20,039 of 25,691 trials) reported summary results within 1 year of primary completion.(10)

While many universities monitor and enforce compliance of reporting for trials with legal reporting requirements, few monitor registration and results reporting that are not legally required. Furthermore, none to our knowledge monitor the reporting of results of trials with principal investigators from other institutions for which their faculty were co-investigators.

This study’s goal was to determine prospective registration, summary results reporting in the registry, publication status, reporting of trial registration numbers in the abstract and in the full text of publications of trials with a primary completion date from 2014 to 2017 with investigators, sponsors, or responsible parties affiliated with seven California universities (in alphabetical order: Stanford University, University of California – Davis, University of California – Irvine, University of California - Los Angeles, University of California - San Diego, University of California - San Francisco, University of Southern California). We chose this period to allow 5-year follow-up for all trials. Our secondary objective was to compare the results with data from German trials with final completion dates from 2014 to 2017,(11) as well as US trials with primary completion dates from 2007 to 2010.(6)

### Methods

Our study was preregistered(12) and based on the methodology used in a prior analysis of trials by German University Medical Centers that are registered in ClinicalTrials.gov as well as the German Clinical Trials Register.(11,13) In short, we identified affiliated trials and obtained their metadata, then searched for trial results publications manually. We then used semi-automatic methods to determine the open access (OA) status of journal publications, time from trial start to its registration and result publication, as well as reporting of trial registration numbers in the abstract or full text of publications. A deviation from our original protocol was the inclusion of two additional data extractors (EZ, NH) as well as a project supervisor (SM), with all three individuals added as co-authors on this publication.

Another addition was the inclusion of trials from University of Southern California (USC). USC was not analyzed in a previous publication that reported data for US trials completed from 2007 to 2010, so we did not include it in the protocol(14). However, we felt that due to the number of trials conducted by USC, it should be included. Additionally, while we wanted to showcase trial funding data and explore it as a possible predictor of practices, details of funding were not available through the metadata sources we used. We did not collect this information manually due to time constraints. Furthermore, although originally planned, statistical comparison of our results with those reported for trials from German University Medical Centers (11) was deemed inappropriate due to differences in the selection criteria (primary completion date in our study vs final completion date in German data). We therefore report that comparison only descriptively in our results section and in the Appendix.

### Outcomes

We aimed to assess (in order presented in the results):

Primary objectives:

a. Timing of trial registration (prospective vs retrospective)
b. Summary results reporting on ClinicalTrials.gov (total, and within 2 and 5 years of trials’ primary completion date)
c. Results reporting in publications (peer reviewed publications, abstract or posters, as well as within 2 and 5 years of trials’ primary completion year)
d. Any form of result reporting (i.e. summary results reporting and results publication described above)
e. Linking of a results publication in the ClinicalTrials.gov registry
f. Trial registration number reporting in the publication abstract and full text
g. Open access status of peer reviewed publications

Secondary objectives:

a. Comparison of results with published data from German trials completed between 2014 to 2017 (11)
b. Comparison of results with published data from US trials from 2007 to 2010 (6)

### Trial Identification and Reporting

We identified trials by using the Clinical Trials Transformation Initiative’s Aggregate Content of ClinicalTrials.gov (AACT) database via the aactr package (RRID:SCR_026245) and searched “overall officials”, “sponsors”, and “responsible parties” fields using name variants and abbreviations of the seven California universities (“sponsors” field was searched only for the sponsor, not the collaborators entries). We then downloaded all such trials and filtered that dataset for trials with a primary completion date (defined as the date that the last data point for the primary outcome measure was collected from the last enrolled participant) from 2014 to 2017. All R (RRID:SCR_001905) scripts used to retrieve, combine, and filter the trials, as well as populate metadata about the trials can be found at the project GitHub (RRID:SCR_002630) repository.(15) We removed duplicates and verified the data with manual checks. In the presentation of summary results, each unique trial was counted only once. In the presentation of trials affiliated with a university, if a trial’s “overall officials”, “sponsors”, or “responsible parties” field had individuals from more than one California University, we counted those trials as affiliated with each of those universities (Figure 1). Specifically, in our sample there were 1,091 unique trials, 1072 (98.3%) were affiliated with only one university, 17 (1.6%) were affiliated with two universities, and 2 (0.1%) were affiliated with three universities.

**Figure 1.**
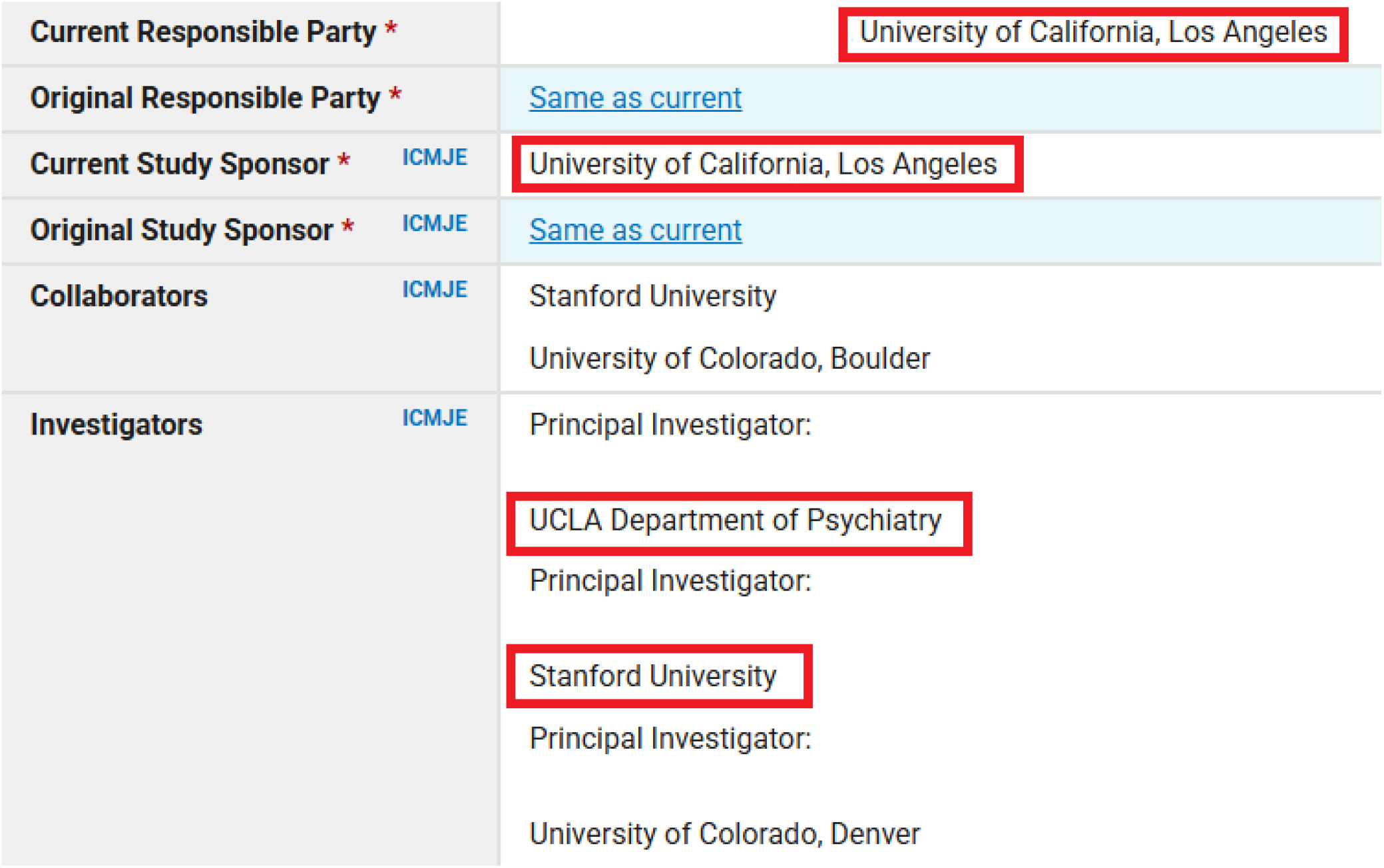
Example of Identification and Classification of a Trial Affiliated with Two California universities.

The figure is a screenshot from the Clinical Trial Registry record <u>NCT01483391</u>, under the category “Administrative information” We removed the names of the individuals, and added red frames around affiliations to California universities identified by our search strategy that searched for names and acronyms of seven California universities. For summary results reporting, this trial was counted once, and when reporting results for trials affiliated with individual universities, it was counted twice, i.e., it was included in results of trial affiliated with Stanford University and with University of California, Los Angeles.

Under FDA and NIH regulations only one university, i.e. commonly the university whose Protocol Registration and Results System (PRS) trial account is used to register the study, holds the legal and contractual responsibility for trial registration and results reporting in the public registry, and is listed as the lead sponsor of the study in ClinicalTrials.gov. However, ClinicalTrials.gov and the related AACT database do not have fields which indicate what trials fall under FDAAA regulations (i.e., Applicable Clinical Trials) or the NIH mandates, nor do they contain a specific PRS Account field, i.e. field that specifies which entity is responsible for reporting trial results. Therefore, the results presented here should not be interpreted as related to legal or funder mandates. Instead, the results should be viewed in relation to ethical recommendations for all parties involved (e.g. Declaration of Helsinki: “*Researchers, authors, sponsors, editors and publishers all have ethical obligations with regard to the publication and dissemination of the results of research*”).(1) Also applicable are the principles of ethical research which specify that the burden on trial participants should be commensurate with a trial’s scientific value, which equal zero if the results are not publicly reported, and are markedly diminished if reporting is not timely.(16) This ethical requirement applies to all trials and trial investigators, not just to the PI, first author, sponsor, or other responsible party.

### Results Publication Search

Data extractors applied a 3-step process for identifying results publications:

1. They first manually checked if a publication of results was linked in the ClinicalTrials.gov registry. If multiple publications were linked, the earliest publication in which primary outcomes were reported was chosen.
2. If the results publication were not linked in the registry, the clinical trial identifier (NCT ID) was entered in Google search, and the first two pages of search results were checked for possible results publications.
3. If results publications were not identified using steps 1 and 2, extractors then used the title of the trial and the name of the principal investigator as a search query in Google. When multiple investigators were listed, individual searches were repeated using the trial title and the name of the other investigator(s). First two pages of results were checked.

When a publication was found, the Digital Object Identifier (DOI) or link and (where available) PMID were extracted for that publication and entered in a database. Date of publication for all trials with a PMID was extracted from PubMed, with detailed steps reported in our Appendix. For those without a PMID, the date of publication was extracted manually. We also extracted data on whether the results were linked in the registry or found through Google search, and what type of results were reported:

A. summary results only (i.e., in the registry),
B. peer reviewed or preprint publication with results, or
C. other (e.g., conference abstract, poster, or presentation slides).

Data extraction was split among six extractors (in alphabetical order by first name: EZ, MM, NH, SY, SG, SW). Extractors were first trained on 30 trials. Raters, on average, identified correct results reporting for 24 out of 30 trials, reaching the target 80% agreement threshold set in the protocol. Additionally, as stated in the protocol, for each trial without identified reporting of results by one of the extractors, a second extractor repeated the full search process. Any discrepancies were resolved by consensus or through consultation with the data extraction lead (SW).

### Determining prospective registration

We used two methods to explore if a trial was prospectively registered:

a. Month approach (reported in the main results) - we considered the trial prospectively registered if it was registered before or in the month of the trial start date. This method was used in previous studies and allows leniency, as 84% of trials in our sample lacked information on which day of the month the registration occurred (i.e., only month and year information was available, see Appendix Extraction of Dates).
b. 21 day approach (reported in the Appendix) - we considered the trial to be prospectively registered if it was registered within 21 days of the start date, as mandated by NIH and FDA regulations.(17) For leniency, dates that lacked information on days were transformed to the end of month (see Appendix Extraction of Dates).

### Determining the time to publication

Publication dates were extracted from 3 sources and the earliest of all three times was used. The 3 sources were: 1) manual extraction during identification of publication; 2) PubMed; and 3) Unpaywall database. For those available through PubMed, we obtained both the publication date and the electronic publication date if it was available (for details see Appendix Extraction of Dates).

### Determining open access status

Open accessibility of trials with results published in peer-reviewed publications was assessed using the Unpaywall API.(18) We counted as openly accessible gold, green, and hybrid publications.

### Statistical Analysis

Summary trial (meta)data is reported as the number and percentage of unique trials (N=1,091) for all categorical characteristics. Trial recruitment size is shown as median and interquartile range (IQR) due to skewed data distributions. Trials that were affiliated with more than one institution were allocated to each institution. Specifically, 19 (1.7%) trials were affiliated with more than one institution; 17 (1.6%) with two institutions, and 2 (0.1%) trials with three institutions, making the total sample size N=1,112 for these analyses. Differences in outcomes between the trials affiliated with different universities were compared using a series of chi-squared tests, and differences between trials based on their primary completion year, using a series of chi-squared tests for trend. Comparison with data from the Chen et al. study, covering US trials from 2007 to 2010 (6), was done with chi-squared tests. All data was analyzed using MedCalc v.22.017 (RRID:SCR_015044) or Microsoft Excel v.2211 (RRID:SCR_016137), and statistical program result outputs are available in our project repository.

## Results

### Trial Characteristics

We identified 1,091 unique trials affiliated with seven California universities registered on the ClinicalTrials.gov trial registry that had a primary completion date between 2014 and 2017 (Table 1). All trials were interventional, the majority were no-phase (e.g., studies of devices or behavioral interventions, n=589, 54%), followed by phase 2 studies (n=159, 15%, Appendix Table 1). Most trials were single-center (n=752, 69%) and had a median recruitment size of 50 participants (IQR 21 to 150). The majority had the status completed (n=921, 84%), followed by terminated (n=154, 14%), active – not recruiting (n=11, 1%), and suspended (n=5, 0%).

**Table 1.** Prospective registration, (summary) results reporting and open accessibility of results for clinical trials with primary completion year from 2014 to 2017 affiliated with seven California universities.

| Investigator, sponsor or responsible party affiliation (n, %)<br>(Affiliation does not reflect legal responsibility)* | Preregistered | Summary results in ClinicalTrials.gov | Journal publication | Abstract/Poster | Open Access*** |
| --- | --- | --- | --- | --- | --- |
| Stanford University (N=266) | 162 (61) | 160 (60) | 173 (65) | 4 (2) | 120 (69) |
| UC Davis (N=110) | 60 (55) | 37 (34) | 82 (75) | 4 (4) | 61 (74) |
| UC Irvine (N=41) | 12 (29) | 25 (61) | 23 (56) | 3 (7) | 15 (65) |
| UC Los Angeles (N=188) | 136 (72) | 74 (39) | 128 (68) | 2 (1) | 107 (84) |
| UC San Diego (N=127) | 85 (67) | 68 (54) | 83 (65) | 5 (4) | 68 (82) |
| UC San Francisco (N=299) | 220 (74) | 117 (39) | 228 (76) | 13 (4) | 190 (83) |
| University of Southern California (N=81) | 40 (49) | 28 (35) | 52 (64) | 5 (6) | 35 (67) |
| Total (N=1112)** | 715 (64) | 509 (46) | 769 (69) | 36 (3) | 596 (78) |

\*We defined a trial as affiliated with a university if the university or one of its investigators was mentioned in one of the following fields of the ClinicalTrials.gov registry entries "Overall Official," "Responsible Parties," or "Sponsors." See Methods for more details.
\*\*Trials affiliated with more than one university were counted for each university (i.e., there were 1091 unique trials, of which 1072 were classified as affiliated with 1 California university, 17 with two California universities, and 2 with 3 California universities).
\*\*\*Counted for open access are gold, green, and hybrid versions of journal publications. Denominator for this column was the number of trials with journal publications, while all other columns were based on the total number of trials.

### Trial Registration

Overall, 64% of trials (n=698) were prospectively registered. Prospective registration statistically differed between trials affiliated with different universities (P<0.0001, chi-square test, Table 1), and per trial primary completion year, increasing from 52% for trials with primary completion in 2014 to 71% in those with primary completion date in 2017 (P<0.0001, chi-square for trend). Additional details on prospective trial registration are presented in the Appendix Trial Registration section.

### Summary Results Reporting in the ClinicalTrials.gov Registry

Summary results were reported on ClinicalTrials.gov for 29% of trials (n=313) within 2 years of their primary completion date, and for 44% of trials (n=475) within 5 years. This increased to 46% (n=500) in a period of more than 5 years after completion (Appendix Table 2). Summary results reporting statistically differed among trials affiliated with different universities (P<0.0001, chi-square test, Table 1), and per trial primary completion year (i.e., 2014 to 2017, P=0.0097, chi-square for trend).

**Table 2.** Comparison with German trials registered in ClinicalTrials.gov.

| **Trials (n, %)** | **Preregistered** | **Summary results in ClinicalTrials.gov** | **Journal publication** | **Open Access\*** |
| --- | --- | --- | --- | --- |
| **Germany (2014 to 2017, N=1,213)** | 681 (56)* | 93 (8) | 794 (65) | 454 (59)** |
| **California (2014 to 2017, N=1,091)** | 698 (64) | 500 (46) | 750 (69) | 579 (77) |

\*Sample 1,211 – Two trials had no start date in the database.
\*\*Sample Denominator was 765, as those publications had Unpaywall information

### Results Reported in Publications

For 46% (n=507) of trials, publication (e.g., article, abstract or poster) occurred within 2 years of the primary completion date, and for 69% (n=754) within 5 years (Appendix Table 3). This increased to 72% (n=786) in more than 5 years after completion, with 69% (n=750) of trials published as journal peer-reviewed articles, and an additional 3% (n=36) as only abstracts or posters. Publication rates statistically differed among trials affiliated with different universities (P=0.0026, chi-square test, Table 1), with no statistically significant differences per primary completion year (P=0.6929 chi-square test for trend).

**Table 3.**
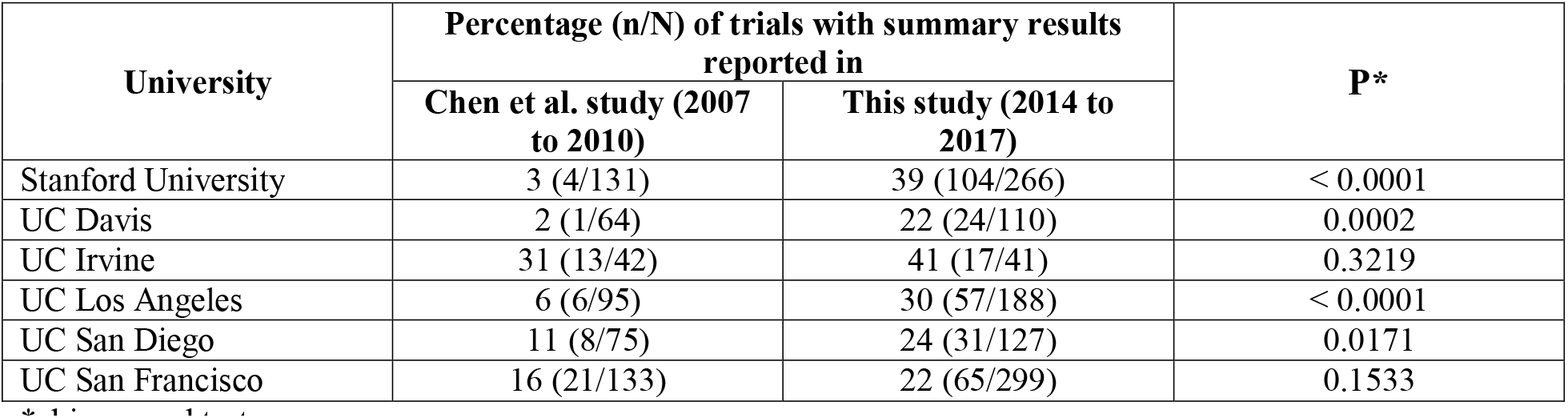
Comparison of summary results reported in ClinicalTrials.gov within 2 years for trials with primary completion year between 2007 and 2010 and those between 2014 to 2017.

### Any Form of Results Reporting

Primary results were reported in any form within 2 years of trials’ primary completion date for 58% (n=637) of trials, and for 81% (n=888) of trials within 5 years. This increased to 84% (n=919) in a period of more than 5 years after completion. Publication rates did not statistically differ among trials affiliated with different universities (P=0.1146, chi-square test, Appendix Table 4), nor per primary completion year (P=0.0773 chi-square test for trend). For the 14% (n=154) of trials listed as terminated early, 49% (n=76) had results disseminated within 2 years, and 68% (n=105) within 5 years.

**Table 4.**
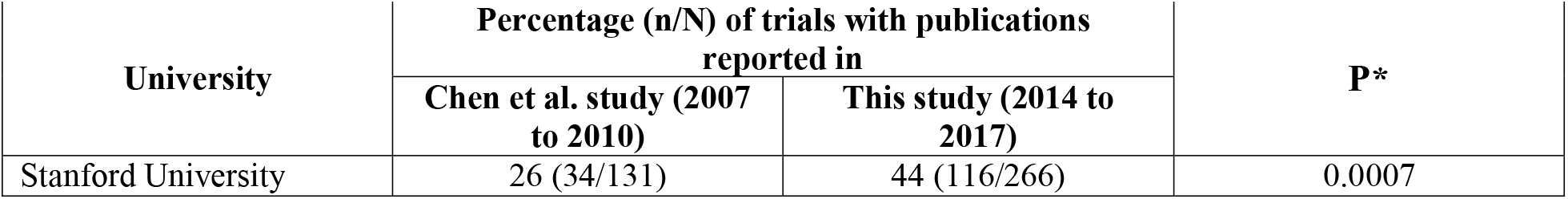
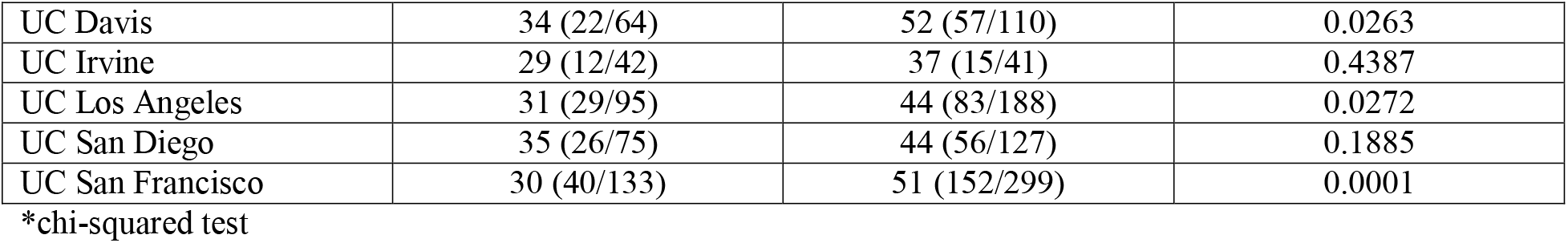
Comparison of results reported in publications within 2 years for trials with primary completion year between 2007 and 2010 and those between 2014 to 2017.

| **University** | **Percentage (n/N) of trials with publications reported in** |  | **P\*** |
| --- | --- | --- | --- |
|  | **Chen et al. study (2007 to 2010)** | **This study (2014 to 2017)** |  |
| Stanford University | 26 (34/131) | 44 (116/266) | 0.0007 |
| UC Davis | 34 (22/64) | 52 (57/110) | 0.0263 |
| UC Irvine | 29 (12/42) | 37 (15/41) | 0.4387 |
| UC Los Angeles | 31 (29/95) | 44 (83/188) | 0.0272 |
| UC San Diego | 35 (26/75) | 44 (56/127) | 0.1885 |
| UC San Francisco | 30 (40/133) | 51 (152/299) | 0.0001 |

\*chi-squared test

### Linked Publications and Reporting of Trial Registration Numbers

Only 58% (n=438) of the 750 trials with results published as peer-reviewed journal articles linked to the article in the trial registry. Linking to the articles in the registry did not statistically differ among trials affiliated with different universities (P=0.5188, chi-square test, Appendix Table 5), nor per study primary completion year (P=0.900, chi-square test for trend).

**Table 5.** Comparison of any form of results reporting for trials with primary completion year between 2017 and 2010 and those between 2014 to 2017.

| University | Percentage (n/N) of trials with any form of results reporting reported in |  | P* |
| --- | --- | --- | --- |
|  | Chen et al. study (2007 to 2010) | This study (2014 to 2017) |  |
| Stanford University | 54 (71/131) | 85 (227/266) | < 0.0001 |
| UC Davis | 69 (44/64) | 84 (92/110) | 0.0223 |
| UC Irvine | 62 (26/42) | 88 (36/41) | 0.0070 |
| UC Los Angeles | 68 (65/95) | 78 (147/188) | 0.0739 |
| UC San Diego | 63 (47/75) | 86 (109/127) | 0.0002 |
| UC San Francisco | 74 (99/133) | 88 (263/299) | 0.0004 |

\*chi-squared test

The trial registration number was listed in the abstract of the associated results publication in 37% (n=276) of articles, and in the manuscript (or declarations) in 45% (n=336) of articles. Trial registration number reporting in abstracts statistically differed among trials affiliated with different universities (P=0.0027, chi-square test) and per study primary completion year (P=0.0401 chi-square test for trend).

Trial registration number reporting in manuscripts did not statistically differ among trials affiliated with different universities (P=0.2399, chi-square test), nor per study primary completion year (P=0.2695, chi-square test for trend).

### Open Access status of Results Publications

Out of the 750 trials with results published as peer-reviewed journal articles, 77% (n=579) were published in open access journals, of which 48% (n=360) were green open access, 22% (n=163) gold, and 7% (n=56) hybrid. Open access rates statistically differed among trials affiliated with different universities (P=0.0025, chi-square test, Table 1), and per primary completion year (P=0.0397, chi-square test for trend).

### Comparison with German Trials from 2014 to 2017

In Table 2, we display these data with those from German university medical centers. As the present sample included trials with primary completion dates (last day of data collection for primary study outcomes) from 2014 to 2017, while the German sample included trials with final completion dates (last day of data collection for all study outcomes) from 2014 to 2017, no statistical comparisons were made.

### Comparison within USA

Chen et al. included data from California trials with primary completion dates from October 2007 to September 2010 that were affiliated with academic medical centers that in that period had more than 40 trials.(6) Results were searched from January to July 2014. Overall, 2,892 (67%) of the 4,347 clinical trials had been published or reported results as of July 2014, of which 1,560 (36%) trials had been disseminated within 24 months. Direct comparisons of data from California universities are shown in Table 3 for summary results reporting in the registry within 2 years, in Table 4 for results reported in publications within 2 years, and in Table 5 for any form of results reporting (Note: University of Southern California was not analysed in Chen et al.).

## Discussion

Our study has shown that about one fifth (19%) of 1,091 trials with investigators, sponsors, or responsible parties affiliated with seven California universities with primary completion dates from 2014 to 2017 lacked any kind of results dissemination (e.g., summary results in registry, journal article, poster or abstract) 5 years after their primary completion date. While 58% of trials had results disseminated within 2 years, less than half (46%) reported summary results on ClinicalTrials.gov registry. These results are similar to those recently published for a random sample of 400 US trials initiated after 2015 with primary completion dates prior to 2018, for which 48% reported summary results. (19). While summary results reporting in our sample is higher than that reported for Canadian(20), German(11), or Polish trials(21) registered on ClinicalTrials.gov registry, they are still lower than for trials covered by the FDAAA (or NIH) mandates (77% reporting of summary results in the registry)(10), indicating that such mandates, even when not imposing or enforcing financial penalties for non-reporting, seem to increase trial summary results reporting.

Furthermore, our findings of clinical trial registration numbers being reported in 37% of publication abstracts, and in 45% of manuscripts, is slightly lower than that reported for German trials (38% and 60%, respectively)(13), and in a random sample of 200 USA trials, where trial number linkage in the abstract was present for 55% of trials with results publications (22). The open access status of publications in our study was 77%, which was higher than 59% of German trials (though that also included bronze as open access).(24)

International guidelines such as the Declaration of Helsinki (1) and WHO Joint Statement(2) provide clear ethical recommendations for prospective trial registration and results reporting, with WHO specifying timely dissemination as summary results reporting within 12 months and journal publication within 24 months after completion. However, legal and institutional requirements around the world vary as to whether and how they regulate or enforce these recommendations. To ensure compliance with both ethical principles and regulatory requirements, universities need effective monitoring and support systems.

This study has several limitations and important caveats. First, as we employed manual and semi-automatic searches and did not contact principal investigators directly, it is possible that some results reporting was missed or that we did not pick the earliest results reporting study. In a follow-up of the German study, 6% of trial publications were either not identified or wrongly identified based on the PI feedback. This did not affect the 2-year and 5-year publication rates in that sample.(23)

Second, we included all trials in results reporting, even those with terminated status, as 33% of those in our sample reported results. Some terminated trials may not have recruited any participants and their registrations were not updated to a withdrawn status. Other terminated trials may have recruited so few participants that results would be uninformative, although the results reporting rate of such trials were only modestly lower than for other trials.

Third, while we examined differences between the California universities, and between trials with different primary completion years, any difference among them may be due to trial characteristics that were not captured in our study, such as characteristics of researchers.

Finally, we repeat the caveat that these results should not be interpreted as related to legal or funder mandates, because analysis of legal liability was not our objective. Our study included trials with primary completion dates between 2014 and 2017, while the FDAAA 2007 Final Rule came into force in January 2017. According to the FDAAA Trials Tracker, only 92 (8%) of trials in our study are subject to the FDAAA mandate. Of these 92, 65 (71%) were reported late, and 9 (10%) were still overdue to report their results in January 2025. Of the 9 overdue, only 2 have California Universities as the PRS account holders, i.e. as responsible parties for reporting of results. So of those trials that were legally required to report results, only 2.1% (2/92) were overdue and the primary responsibility of a California university to report.

In conclusion, our study has shown that timely results reporting may be improving, but for studies completed in 2014-2017, most of which were not subject to FDA or NIH reporting requirements and mandates, 2-year results are suboptimal, and even 5-year results reporting has substantial room for improvement.

## Data Availability

The initial registry data from the AACT is available at https://github.com/maia-sh/california-trials. The final dataset used for analysis is available at https://github.com/ontogenerator/california-clinical-dashboard/tree/main/data.

https://doi.org/10.25740/gq169sj2027

## Acknowledgements

None.

