## Appendix for "Registration and Reporting of Clinical Trials Affiliated with California Universities and with Primary Completion Date from 2014 to 2017"

Results are presented in the order they appear in the manuscript. All data was explored using MedCalc v.20.123 (RRID:SCR_015044) or Microsoft Excel v.2211 (RRID:SCR_016137).

### Inter-rater agreement

While initially 3 raters had the slightly lower results than prespecified threshold (2 77%, and one 67%), we included all in the extraction process, as raters mistakes were related to trials having more than one publication with results, and raters not choosing the earliest one - and not making a mistake of stating there was no publication when there was one. Therefore, the final results, might slightly impact the time to publication rather than the rates of publications.

Extraction of Dates

Trial start date, registration date and primary completion year date were extracted from the Clinical Trials Transformation Initiative’s Aggregate Content of ClinicalTrials.gov (AACT) database. Majority of trials did not contain information on day of the month.

| **Variable** | **n (%)** |
| --- | --- |
| Study start date | 913 (84) |
| Study primary completion date | 765 (70) |
| Publication date | 7 (1) |

Publication dates information came from 3 sources: manual extraction, PubMed, or Unpaywall data. Dates from PubMed were retrieved in the following way: we manually copied the list of all publications PMIDs (separated with a blank space) as a search strategy in PubMed, and then exported the results as csv file. From the citation variable of the csv export, we then extracted the publication dates and the e-publication dates (if available).

If a paper had only a manually extracted publication date, that date was used for all date comparisons. If a publication also had dates retrieved from Unpaywall and PubMed, we took the earliest date associated with it as the publication date (either electronic or print publication date from PubMed or the date given on Unpaywall). Publication dates listed on Unpaywall without the exact day of the month were disregarded.

### Results

Appendix Table 1. **Phase type of trials (n, %):**

| Early Phase 1 | 25 | 2% |
| --- | --- | --- |
| Not Applicable | 589 | 54% |
| Phase 1 | 103 | 9% |
| Phase 1/Phase 2 | 54 | 5% |
| Phase 2 | 159 | 15% |
| Phase 2/Phase 3 | 18 | 2% |
| Phase 3 | 64 | 6% |
| Phase 4 | 79 | 7% |
| Total | 1091 | 100.0% |

Study Dates

CT.gov allowed entering only year and month for dates related to study start, registration, and completion. We obtained these three dates from AACT database. 84% of studies had month/year dates for study start, 70% for primary completion date, and 68% for completion date with sig. differences between the universities, as well as primary completion years, and study start years for all 3 of those dates (data not shown).

If we look at trials for which we have the exact dates for study start (N=178), there were 56% prospectively registered (when prospectively registered is calculated as being the same day of before the study start date), when we take into consideration the USA law (21 days after start day) – 63% are “registered on time”.

Trial Registration

The results for prospective registration presented in the manuscript results were calculated using the month method (see details in manuscript methods). As an additional analysis, we also calculated the time to registration taking into the account 21 days following the study start date (see methods). Those results are presented here:

Overall, 59% of trials (n=641) were prospectively registered. Prospective registration statistically differed between Universities (P<0.0001, chi-square test, Table 1), and per trial primary completion year, growing from 49% for trials with primary completion in 2014 to 66% in those with primary completion date in 2017 (P<0.0001, chi-square for trend).

Appendix Table 2. **Summary Results reported in ClinicalTrials.gov registry for trials with primary completion date from 2014 to 2017 affiliated with seven California universities.**

| **Investigator, sponsor or responsible party affiliation (n, %)**  **(Affiliation does not reflect legal responsibility)*** | **Summary results in CT.gov** | **Summary results in CT.gov within 2 years** | **Summary results in CT.gov within 5 years** |
| --- | --- | --- | --- |
| Stanford University (N=266) | 160 (60) | 104 (39) | 157 (59) |
| UC Davis (N=110) | 37 (34) | 24 (22) | 37 (34) |
| UC Irvine (N=41) | 25 (61) | 17 (41) | 23 (56) |
| UC Los Angeles (N=188) | 74 (39) | 57 (30) | 73 (39) |
| UC San Diego (N=127) | 68 (54) | 31 (24) | 61 (48) |
| UC San Francisco (N=299) | 117 (39) | 65 (22) | 106 (35) |
| University of Southern California (N=81) | 28 (35) | 21 (26) | 27 (33) |
| Total (N=1112)** | 509 (46) | 319 (29) | 484 (44) |
| P (chi-squared test) | < 0.0001 | 0.0001 | < 0.0001 |

*We defined a trial as affiliated with a university if the university or one of its investigators was mentioned in one of the following fields of a clinicaltrials.gov registry entries "Overall Official," "Responsible Parties," or "Sponsors." See Methods for more details.

**Trials involving individuals from more than one university were counted for each university (i.e., there were 1091 unique trials, of which 1072 were classified as involving 1 California university, 17 involving two California universities, and 2 involving 3 California universities).

Appendix Table 3. **Publications rate for trials with primary completion date from 2014 to 2017 affiliated with seven California universities.**

| **Investigator, sponsor or responsible party affiliation (n, %)**  **(Affiliation does not reflect legal responsibility)*** | **Journal publication** | **Abstract/Poster** | **Publication within 2 years** | **Publication within 5 years** | **Open Access**** |
| --- | --- | --- | --- | --- | --- |
| Stanford University (N=266) | 173 (65) | 4 (2) | 116 (44) | 172 (65) | 120 (69) |
| UC Davis (N=110) | 82 (75) | 4 (4) | 57 (52) | 81 (74) | 61 (74) |
| UC Irvine (N=41) | 23 (56) | 3 (7) | 15 (37) | 25 (61) | 15 (65) |
| UC Los Angeles (N=188) | 128 (68) | 2 (1) | 83 (44) | 125 (66) | 107 (84) |
| UC San Diego (N=127) | 83 (65) | 5 (4) | 56 (44) | 85 (67) | 68 (82) |
| UC San Francisco (N=299) | 228 (76) | 13 (4) | 152 (51) | 231 (77) | 190 (83) |
| University of Southern California (N=81) | 52 (64) | 5 (6) | 38 (48) | 53 (65) | 35 (67) |
| Total (N=1112)*** | 769 (69) | 36 (3) | 518 (47) | 772 (69) | 596 (78) |
| P (chi-squared test) | 0.0026 | | 0.3377 | 0.0165 | 0.0025 |

*We defined a trial as affiliated with a university if the university or one of its investigators was mentioned in one of the following fields of a clinicaltrials.gov registry entries "Overall Official," "Responsible Parties," or "Sponsors." See Methods for more details.

**Counted for open access are gold, green, and hybrid versions of journal publications. Denominator for this column was the number of trials with journal publications, while all other columns were based on the total number of trials.

**Trials involving individuals from more than one university were counted for each university (i.e., there were 1091 unique trials, of which 1072 were classified as involving 1 California university, 17 involving two California universities, and 2 involving 3 California universities).

Appendix Table 4. **Any Results Dissemination for trials with primary completion date from 2014 to 2017 affiliated with seven California universities.**

| **Investigator, sponsor or responsible party affiliation (n, %)**  **(Affiliation does not reflect legal responsibility)*** | **Any Result Dissemination** | **Any Result Dissemination within 2 years** | **Any Result Dissemination within 5 years** |
| --- | --- | --- | --- |
| Stanford University (N=266) | 227 (85) | 159 (60) | 224 (84) |
| UC Davis (N=110) | 92 (84) | 62 (56) | 87 (79) |
| UC Irvine (N=41) | 36 (88) | 24 (59) | 34 (83) |
| UC Los Angeles (N=188) | 147 (78) | 105 (56) | 141 (75) |
| UC San Diego (N=127) | 109 (86) | 73 (57) | 104 (82) |
| UC San Francisco (N=299) | 263 (88) | 178 (60) | 255 (85) |
| University of Southern California (N=81) | 65 (80) | 50 (62) | 62 (77) |
| Total (N=1112)** | 939 (84) | 651 (59) | 907 (82) |
| P (chi-squared test) | 0.1146 | 0.9620 | 0.0811 |

*We defined a trial as affiliated with a university if the university or one of its investigators was mentioned in one of the following fields of a clinicaltrials.gov registry entries "Overall Official," "Responsible Parties," or "Sponsors." See Methods for more details.

**Trials involving individuals from more than one university were counted for each university (i.e., there were 1091 unique trials, of which 1072 were classified as involving 1 California university, 17 involving two California universities, and 2 involving 3 California universities).

Appendix Table 5. **Publication links and trial registration number reporting for trials with primary completion date from 2014 to 2017 affiliated with seven California universities.**

| **Investigator, sponsor or responsible party affiliation (n, %)**  **(Affiliation does not reflect legal responsibility)*** | **Journal publication** | **Publications linked in CT.gov** | **Trial Registration Number in Abstract** | **Trial Registration Number in Publication** |
| --- | --- | --- | --- | --- |
| Stanford University (N=266) | 173 (65) | 98 (57) | 44 (25) | 73 (42) |
| UC Davis (N=110) | 82 (75) | 48 (59) | 37 (45) | 41 (50) |
| UC Irvine (N=41) | 23 (56) | 11 (48) | 5 (22) | 6 (26) |
| UC Los Angeles (N=188) | 128 (68) | 81 (63) | 54 (42) | 59 (46) |
| UC San Diego (N=127) | 83 (65) | 50 (60) | 37 (45) | 31 (37) |
| UC San Francisco (N=299) | 228 (76) | 137 (60) | 92 (40) | 111 (49) |
| University of Southern California (N=81) | 52 (64) | 25 (48) | 16 (31) | 23 (44) |
| Total (N=1112)** | 769 (69) | 450 (59) | 285 (37) | 344 (45) |
| P (chi-squared test) | 0.0026 | 0.5188 | 0.0027 | 0.2399 |

*We defined a trial as affiliated with a university if the university or one of its investigators was mentioned in one of the following fields of a clinicaltrials.gov registry entries "Overall Official," "Responsible Parties," or "Sponsors." See Methods for more details.

**Trials involving individuals from more than one university were counted for each university (i.e., there were 1091 unique trials, of which 1072 were classified as involving 1 California university, 17 involving two California universities, and 2 involving 3 California universities).
